# Inflow restrictions can prevent epidemics when contact tracing efforts are effective but have limited capacity

**DOI:** 10.1101/2020.04.01.20050401

**Authors:** Hannes Malmberg, Tom Britton

**Affiliations:** Department of Economics, University of Minnesota; Department of Mathematics, Stockholm University

## Abstract

When a region tries to prevent an outbreak of an epidemic, like that of COVID-19, two broad strategies are initially available: limiting the inflow of infected cases using travel restrictions and quarantines, and reducing the transmissions from inflowing cases using contact tracing and community interventions. A large number of papers have used epidemiological models to argue that inflow restrictions are unlikely to be effective. We conduct a mathematical analysis using a simple epidemiological model and perform simulations which show how this conclusion changes if we relax the assumption of unlimited capacity in containment efforts such as contact tracing. In particular, when contact tracing is effective, but the system is close to being overwhelmed, moderate travel restrictions can have a very large effect on the probability of an epidemic.

## 1. Introduction

Two main factors determine whether and when a region will be affected by an epidemic like the current COVID-19 outbreak. The first factor is the rate *λ* at which infectious individuals (either visitors or returning local residents) enter the country. The second factor is the probability *π* that such an entry gives rise to a major outbreak.^1^

Potential preventive measures by health authorities can hence target reduction of *λ*, by e.g. travel restrictions or quarantines, or reduction of *π*, by e.g. contact tracing in conjunction with isolating the imported cases (henceforth “contact tracing”). Quite often preventive measures aim at reducing both of these factors.

Contact tracing can be fully effective (i.e., *π* = 0) if it manages to bring the epidemic’s effective reproduction number *R* below 1 during the early stage of the outbreak, where *R* is defined as the expected number of infections caused by a typical infected individual. Intuitively, a large outbreak is not possible if each infected individual, on average, spreads the disease to less than one other individual.

We use a simple epidemic model to analyze the effects of reducing *π* and *λ* on epidemic outbreaks. The background to our work is a literature that has found small effects from regulating *λ*. Anzai *et al*. show that inflow reduction alone (assuming *π* > 0) cannot prevent an epidemic outbreak from taking place and at best delays epidemic onset, often for just a very limited time (*1*). T Chinazzi *et al*. study the effect of reducing the inflow of infected individuals while simultaneously reducing *π* in the community at large and also find that inflow reduction has only a marginal delay effect unless *π* is reduced drastically (2). These pessimistic findings reflect those from a large number of earlier models (*3-8*). The World Health Organization’s 2014 Systematic Review on the role of travel restrictions in containing pandemic influenza reviewed 23 papers and concluded that a 90% reduction in international air travel would only slow down a pandemic by 3-4 weeks and would not prevent the introduction of a pandemic into any given country *(9*).

We focus on the joint effect of inflow reduction and contact tracing. Contact tracing is incorporated in our model by having two different outbreak probabilities, one negligible for incoming cases that are contact traced, and another much higher probability for incoming cases that are not contact traced. If no one is contact traced, then reductions in *λ* have only very modest effects, in line with the previous results mentioned above. Furthermore, if every infected inflow is contact traced and contact tracing is fully effective, then changing the inflow rate *λ* is still ineffectual, since there will never be an epidemic outbreak. Hence, in the extreme cases where either no one or everyone is contact traced, changing *λ* does not meaningfully affect the probability of an outbreak.

However, this conclusion relies critically on there being unlimited capacity in contact tracing, i.e., that all imported cases are contact traced nearly flawlessly irrespective of how many are currently being traced. We argue that when contact tracing has limited capacity but is still effective, reducing *λ* may well be very effective in reducing the risk of an outbreak. Regulating *λ* is particularly important when the contact tracing system is close to being overwhelmed by new cases arriving from outside, in which case even moderate reductions in *λ* can strongly reduce the probability of an epidemic outbreak.

Since contact tracing is both resource- and labor-intensive, we believe that our limited capacity assumption is reasonable. We also conjecture that, while our model is simple, our qualitative findings will translate to more realistic setups. Hence, we think that epidemiological models used for policy analysis should incorporate capacity constraints, since they might otherwise underestimate the potential of travel restrictions to prevent epidemic outbreaks (or the re-emergence of an epidemic).

## 2. Model

### 2.1 Setup

Formally, we study a stochastic epidemic model where infected cases arrive at some Poisson rate *λ*. Absent contact tracing, each new case leads to an epidemic outbreak with some probability *π*_*NT*_ > 0. The containment effort consists of contact tracing that reduces the probability of an epidemic outbreak to *π*_*T*_ ≥ 0. We say that contact tracing is *effective* if *π*_*T*_ = 0 and is *imperfect* if *π*_*T*_ > 0.

To model potential capacity constraints, we assume that contact tracing is conducted by a set of *n* teams which process every case arrival with an intensity *μ* (so the mean time for completing contact tracing is 1/*μ*). If a case arrives when at least one team is free, that case has probability *π*_*T*_ of leading to an epidemic. If all teams are occupied when an infected case arrives, that case is lost to the system and causes an epidemic with probability *π*_*NT*_. We say that contact tracing has *unlimited capacity* if *n* = ∞ and has *limited capacity* if *n* < ∞.

This setup can be modelled as a queuing system where the state is the number of people currently in the contact tracing system. To analyze the probability of an epidemic outbreak at different time horizons, we add the outbreak as an additional, absorbing state. The setup is illustrated in Fig. 1.

**Fig. 1.**
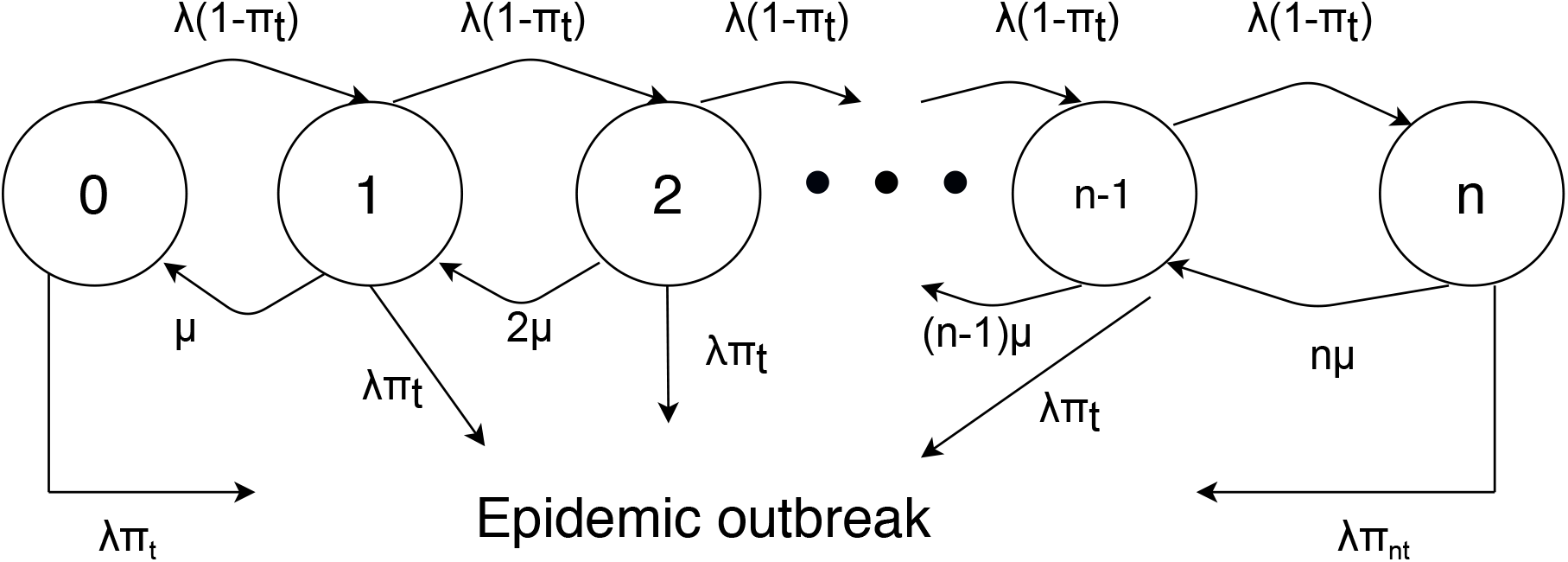
Simplified model of epidemic outbreak with contact tracing. This diagram outlines the basic evolution of a disease from emergence to epidemic outbreak in the presence of a contact tracing system. When the system is imperfect, each traced case has a positive probability of leading to an epidemic, regardless of the arrival rate of new cases, the rate at which cases are processed, or the number of cases that can be processed at once. When the system is effective, an outbreak will only occur if the system’s capacity is limited and not all newly arriving cases can be processed.

### 2.2 The effect of contact tracing on outbreak probability

With this setup, we consider the effect of varying *λ* under four different combinations of cases: contact tracing being imperfect versus effective and having unlimited versus limited capacity.

Doing this, we confirm the recent literature’s main finding: varying *λ* is relatively inconsequential when capacity is unlimited. We also find, however, that varying *λ* can be very important when contact tracing is effective but has limited capacity. We proceed by discussing each case in turn.^2^

#### Case 1 – Imperfect contract tracing with unlimited capacity

In this case, there is an epidemic outbreak at a constant rate *λπ*_*T*_ > 0. At a horizon *t*, the probability of an outbreak is

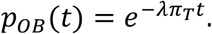

Reducing *λ* can proportionally delay the outbreak but cannot stop it.

#### Case 2 – Effective contact tracing with unlimited capacity

In this case, all arriving cases have zero probability of causing an epidemic. Thus, regardless of *λ*, there will not be an outbreak.

#### Case 3 – Imperfect contact tracing with limited capacity

In this case, epidemics break out at a rate *λπ*_*T*_ when the contact tracing system is below capacity and at a rate *λπ*_*NT*_ > *λπ*_*T*_ when at capacity. In contrast with Case 1, reducing *λ* has the benefit of reducing the probability of being at full capacity. However, since the containment system is not fully effective, reducing *λ* can still only delay the outbreak.

#### Case 4 – Effective contact tracing with limited capacity

In this case, epidemics do not occur when the contact tracing system is below capacity but do occur at a rate *λπ*_*NT*_ > 0 when at capacity. Thus, by preventing the system from reaching full capacity, reducing *λ* can be very effective at preventing outbreaks. The effect of *λ* can also be highly non-linear. Indeed, unless the queue was truncated at *n*, the value *λ* = *nμ* would be a critical value where the system discretely would change from being a subcritical to a supercritical system.^3^

## 3. Simulation

To illustrate our findings, we perform simulations varying *λ* under each of these four different cases. All simulation parameters are given in Table 1. We assume that when capacity is limited, there are *n* = 100 tracing teams and cases are handled by a team, on average, in two days (i.e., *μ* = 0.5). We consider three infected case arrival rates: a baseline rate *λ* = 80, a moderate reduction *λ* = 40, and a strong reduction *λ* = 20.^4^ When contact tracing is imperfect, we assume that *π*_*T*_ = 0.1%, and when contact tracing is effective, we assume that that *π*_*T*_ = 0.001%.^5^ In the absence of any contact tracing, we assume that *π*_*NT*_ = 10%.

**Table 1.** This table lists all parameter values used in our model simulated experiments. For parameters where multiple values are considered, trailing parentheticals indicate each value’s relevant case.

| Parameter | Value(s) |
| --- | --- |
| $n$ | 100 (*limited*), ∞ (*unlimited*) |
| $\mu$ | 0.5 |
| $\lambda$ | 80 (*baseline*), 40 (*moderate*), 20 (*strong*) |
| $\pi_T$ | $10^{-3}$ (*imperfect*), $10^{-5}$ (*effective*) |
| $\pi_{NT}$ | 0.1 |

The results for each case are displayed in Fig. 2, with the number of days on the horizontal axis and the probability of epidemic outbreak on the vertical axis. The first row shows the first two cases, where contact tracing has unlimited capacity. In the left-hand panel, contact tracing is imperfect, and reducing *λ* only delays the outbreak. In the right-hand panel, contact tracing is effective and there is a low probability of an outbreak independent of *λ*.

**Fig. 2.**
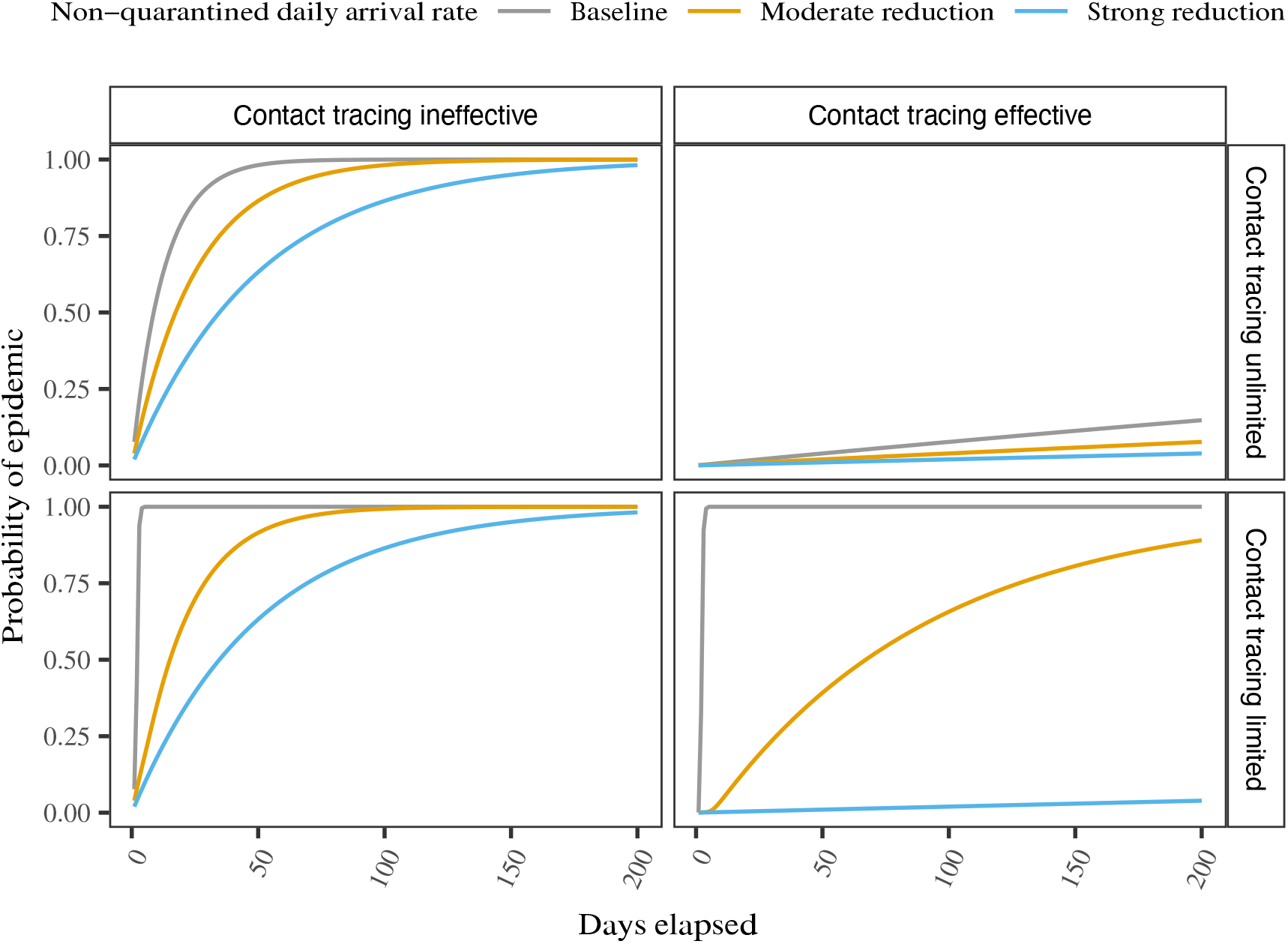
Model-simulated probability of an epidemic outbreak as a function of time. In each panel are the results of model simulations estimating the probability of an epidemic occurring as a function of time given one of three possible arrival rates for new infected cases: a baseline arrival rate, a moderately reduced arrival rate, and a strongly reduced arrival rate. The top row of panels displays results from simulations where contact tracing is assumed to have unlimited capacity, while the bottom row assumes limited capacity. The left column of panels displays results from simulations where contract tracing is assumed to be less than fully effective, while the right column assumes it is fully effective.

The second row shows the two cases where contract tracing has limited capacity. In the left-hand panel, contact tracing is imperfect and the result is qualitatively similar to the case with unlimited capacity; reducing *λ* only delays the epidemic, with a somewhat stronger effect in that the epidemic starts immediately in the absence of inflow restrictions. In contrast, when contact tracing is effective, the result is very different than with unlimited capacity. This case is shown in the right-hand panel, where reducing *λ* can change the probability of an epidemic from a virtual certainty to almost nil by preventing the tracing system from being overwhelmed.

Taking stock, we conclude that introducing capacity constraints can imply large changes in the effectiveness of inflow restrictions. Instead of inflow restrictions leading to a gradual delay of an epidemic, there are non-linear effects once the system goes below capacity. Thus, in cases where systems are at the risk of being overrun, even moderate travel restrictions can be highly effective in reducing the risk for a local epidemic.

While most rich countries are now beyond the point of preventing domestic outbreaks of COVID-19, we believe that the reasoning in this paper is still relevant for countries that have outbreaks that are later brought under control. In these cases, inflow restrictions may be helpful in preventing an epidemic from re-emerging, as they allow the country to stay below capacity in their contact tracing efforts.

## Data Availability

Code is available upon request.

## Data and materials availability

Replication code for the simulation exercises is available upon request.

## Footnotes

1 Our analysis focuses on the probability of an epidemic outbreak. We do not study the subsequent evolution of the disease after this event. In general, inflow restrictions become inconsequential once the amount of local transmission is much larger than the number of import cases.

2 All formal derivations are given in the Supplementary Material.

4 These rates are chosen to pass the critical value of *nμ* = 50, the maximum number of cases the teams can handle per day.

5 We use a strictly positive value in order to capture how even a contact tracing system that generally reduces *R* below 1 can still miss an infection chain by a stroke of bad luck.

## References

1. A. Anzai, T. Kobayashi, N. M. Linton, R. Kinoshita, K. Hayashi, A. Suzuki, Y. Yang, S. Jung, T. Miyama, A. R. Akhmetzhanov, H. Nishiura, Assessing the impact of reduced travel on exportation dynamics of novel coronavirus infection (COVID-19). Journal of Clinical Medicine 9(2), 601 (2020).

2. M. Chinazzi, J. T. Davis, M. Ajelli, C. Gioannini, M. Litvinova, S. Merler, A. P. y Piontti, K. Mu, L. Rossi, K. Sun, C. Viboud, X. Xiong, H. Yu, M. E. Halloran, I. M. Longini Jr., A. Vespigani, The effect of travel restrictions on the spread of the 2019 novel coronavirus (COVID-19) outbreak. Science, 10.1126/science.aba9757 (2020).

3. A. L. P. Mateus, H. E. Otete, C. R. Beck, G. P. Dolan, J. S. Nguyen-Van-Tam, Effectiveness of travel restrictions in the rapid containment of human influenza: a systematic review. Bull. World Health Organ. 92(12), 868–880D (2014).

4. E. H. Y. Lam, B. J. Cowling, A. R. Cook, J. Y. T. Wong, M. S. Y. Lau, H. Nishiura, The feasibility of age-specific travel restrictions during influenza pandemics. Theor. Biol. Med. Model. 8, 44 (2011).

5. P. Bajardi, C. Poletto, J. J. Ramasco, M. Tizzoni, V. Colizza, A. Vespignani, Human mobility networks, travel restrictions, and the global spread of 2009 H1N1 pandemic. PLoS ONE 6(1), e16591 (2011).

6. M. L. Ciofi delgi Atti, S. Merler, C. Rizzo, M. Ajelli, M. Massari, P. Manfredi, C. Furlanello, G. S. Tomba, M. Iannelli, Mitigation measures for pandemic influenza in Italy: an individual based model considering different scenarios. PLoS ONE 3(3), e1790 (2008).

7. B. S. Cooper, R. J. Pitman, W. J. Edmunds, N. J. Gay, Delaying the international spread of pandemic influenza. PLoS Med. 3(6), e212 (2006).

8. J. M. Epstein, D. M. Goedecke, F. Yu, R. J. Morris, D. K. Wagener, G. V. Bobashev, Controlling pandemic flu: the value of international air travel restrictions. PLoS ONE 2(5), e401 (2007).

9. M. Eichner, M. Schwehm, N. Wilson, M. G. Baker, Small islands and pandemic influenza: potential benefits and limitations of travel volume reduction as a border control measure. BMC Infectious Diseases 9, 160 (2009).

